# Impact of COVID-19 Attributable Deaths on Longevity, Premature Mortality and DALY: Estimates of USA, Italy, Sweden and Germany

**DOI:** 10.1101/2020.07.06.20147009

**Authors:** Sanjay K Mohanty, Manisha Dubey, Udaya S Mishra, Umakanta Sahoo

**Author notes:** Corresponding Author Sanjay K Mohanty Professor, Department of Fertility Studies, International Institute for Population Sciences, Govandi Station Road, Deonar, Mumbai- 400088, India. **Financial disclosure** No financial disclosures were reported by the authors of this paper.

## Abstract

In a short span of four months, the COVID-19 pandemic has added over 0.4 million deaths worldwide, which are untimely, premature and unwarranted. The USA, Italy, Germany and Sweden are four worst affected countries, accounting to over 40% of COVID-19 deaths globally. The main objective of this study is to examine the impact of COVID-19 attributable deaths on longevity, years of potential life lost (YPLL) and disability adjusted life years (DALY) in USA, Italy, Germany and Sweden. Data from United Nation Population Projection, Statista and centre for disease control and prevention were used in the analyses. Life expectancy, YPLL and DALY were estimated under four scenarios; no COVID-19 deaths, actual number of COVID-19 death as of 22^nd^ May, 2020 and anticipating COVID-19 death share of 6% and 10% respectively. The COVID-19 attributable deaths have lowered the life expectancy by 0.4 years each in USA and Sweden, 0.5 years in Italy and 0.1 years in Germany. The loss of YPLL was 1.5, 0.5, 0.1 and 0.5 million in USA, Italy, Germany and Sweden respectively. The DALY (per 1000 population) due to COVID-19 was 4 in USA, 6 in Italy, 1 each in Germany and Sweden. Compression in life expectancy and increase in YPLL and DALY may intensify further if death continues to soar. COVID-19 has a marked impact on mortality. Reduction in longevity premature mortality and loss of DALY is higher among elderly.

## Introduction

COVID-19 attributable deaths are soaring each day in most of the countries with uncertainties over projected numbers, infection fatality ratio, development of a vaccine and possible end of pandemic. Globally, with over 6 million confirmed infections and additional deaths of over 360 thousand by end of May,2020, the COVID-19 attributable deaths accounts for 1% of total all-cause mortality. If the COVID-19 mortality continues with same pace, the life expectancy would begin to shrink by end of the year though the survival threat is more among the elderly and the chronically ill. Rapid spread of the infection as well as its associated fatality may well be due to novel disease, lack of medical know how, ill-prepared health care system, crowding in urban cities, administrative inefficacy, demographic and social determinants etc.

The case fatality ratio (CFR) is a crude measure of mortality, underestimate the mortality impact of COVID-19. An alternative CFR with 14 days’ delay depicts at least twice higher mortality than CFR [1]. The mortality impact of COVID-19 is higher than many other disease [2]. The standardized metrics such as disability adjusted life years (DALY) and years lost due to disability (YLD) are suggested to infer infection fatality by age [3].

Considerable attempts are made on tracing future trajectories, estimation of infection and fatality rate and risk factors of COVID-19 [4-12]. Demographic structure, co-morbidities and health-care burden explain COVID-19 attributable mortality to some extent [13-15]. Most common observation made as regard COVID-19 fatality is its greater risk among elderly and people with comorbidities including hypertension, diabetes, cardiovascular disease, myocardial injury [4,16-22]. The Diamond Princes cruise ship study of Japan, a standard estimate of infection, estimated the overall case fatality ratio of 2.6% as against the same being 13% among the older aged 70 and above [23].

Inadequate testing and misclassification of deaths by cause underestimate the extent of COVID-19 deaths. In USA, the excess deaths due to pneumonia and influenza raise an apprehension as regard miss-classification of COVID-19 deaths in the absence of adequate testing [24]. In Italy, 54% deaths were attributed to COVID-19 making a case for misclassification of cause of death. The COVID-19 attributable mortality has potential to reduce life expectancy in India and seasonal life expectancy in Italy [25-26]. In United States, 1 million deaths from COVID-19 would increase mortality by one–third and reduction in period life expectancy by 3.9 years in 2020 [27].

Mortality impact of COVID-19 is higher in urban counties and the social determinants are significant predictors of its mortality [28]. High and low fatality due to COVID-19 attributed to density and age structure in terms of elderly in UK [29]. Demographic vulnerability of COVID-19 mortality is lower in younger countries in Sub-Saharan Africa than the industrialized countries [30]. The spread of infection and mortality depends on containment measures, health system response and micro-management of epidemic which may alter reproduction number [31].

By April 2020, the case fatality rate varied from 2.2% in South Korea to 13.0% in Italy. USA, Italy, Sweden and Germany were worst hit countries by the pandemic. By end of May 2020, USA had over 1.8 million confirmed cases and over 106 thousand deaths. About 80% of deaths occurred among adults aged 65 years or more [16]. In Italy, the CFR increased from 4.2% to 13.0% within 43 days and 90% of the change was due to increasing age specific case fatality rates [32]. In Italy, USA and Germany, estimated cases of infections are 6 times, 2 times and 1.2 times higher than the number of confirmed cases, respectively [33].

Existing studies of the pandemic on fatality is limited. Given its rise in intensity it becomes pertinent to gauge impact of COVID-19 attributable mortality on longevity, premature mortality and DALY. This will answer questions like “Would additional deaths due to COVID-19 reduce longevity and increase premature mortality and DALY”.

## Data and Methods

We have analysed four worst affected countries; namely USA, Italy, Sweden and Germany that accounts over 40% of all COVID-19 attributable deaths worldwide. The selection of country is guided by the availability of age-specific infection and mortality data and severity of infection. Estimates are provided under four scenarios; no COVID-infection, COVID-infection as of cut-off date (20^th^ May for USA, 18^th^ May 2020 for Italy and Germany and 22^nd^ may for Sweden) and estimates under 6% and 10% COVID-19 death share. Population and mortality data by age group for 2020 were obtained from the United Nation Population Projection [34]. The total deaths obtained from UN projection are estimated deaths in the absence of COVID-19 infection. The age specific COVID-19 attributable deaths for USA is collected from Centres for Disease Control and Prevention [35] and that for Italy, Germany and Sweden is taken from Statista [36-38]. The total number of confirmed cases and deaths for each country is collected from worldmeter website [39]. We have redistributed the total deaths available from worldmeter as per age distribution of deaths for which age data was available. Under the assumption that the estimated deaths without COVID-19 and deaths due to COVID-19 are mutually exclusive, we have added these deaths to derive age specific death rate (number of deaths per 1000 population). The age specific case fatality ratio (ASCFR) was computed for Italy and Sweden from given data. In case of Germany, the age group of number of infections were not uniform and deaths were available for 0-9, 10-19, 20-49, 50-69 and 70-89. We have redistributed the deaths as per population distribution in 10-year age group. In case of USA, we have used the ASCFR of Diamond Cruise Study that had constant rate (0.2) till age 35 beyond which we have taken the age group close to nearest age group [23].

## Methods

Abridged life tables, estimation of years of potential life lost (YPLL) and disability adjusted life years (DALY) are used in the analyses. Estimates are based on the assumptions that COVID-19 attributable deaths are additional deaths that would have been avoided in absence of COVID-19 infection. The probability of death has been constructed from age specific death rate (ASDR). The 10-year abridged life table is used to estimate the life expectancy and other mortality estimates. Estimates are provided under four scenarios. Scenario 1 provides the deaths as estimated from UN population prospects and labelled as deaths without COVID-19. Scenario 2 considers COVID-19 deaths accounting for 6% of total deaths while scenario 3 would increase the death share to 10% of total deaths by the end of the year. Expected deaths due to COVID-19 are distributed in accordance with the age distribution of COVID-19 as of date. A brief description of YPLL and DALY estimation is given below.

### Years of Potential Life Lost (YPLL)

The YPLL is a summary measure of premature mortality that estimates the average years a person would have lived had he or she not died prematurely. It gives higher weight to the deaths occurring at younger ages and lower weight to the deaths at higher ages [40-41]. YPLL is estimated as:

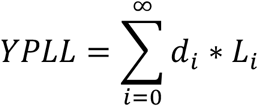

where,

L_i_ is the life expectancy at age i and d_i_ is the number of deaths at age i. The deaths are weighted by life expectancy at each age.

### Disability Adjusted Life Years (DALY)

DALY measures the health of a population by combining data on mortality and non-fatal health outcomes into a single number. The DALY measures health gaps as opposed to health expectancies. It measures the difference between a current situation and an ideal situation where everyone lives up to the age of the standard life expectancy, and in perfect health. It combines in one measure the time lived with disability and the time lost due to premature mortality:

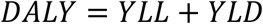

where, YLL= years of life lost due to premature mortality and YLD= years lived with disability.

We have calculated YLL and YLD with discounting rate of 3% where discounting health with time reflects the social preference of a healthy year now, rather than in the future. The value of a year of life is generally decreased annually by a fixed percentage. For many years, a discount rate of 5% per annum has been standard in many economic analyses of health and in other social policy analyses, but recently environmentalists and renewable energy analysts have argued for lower discount rates for social decisions. The World Bank Disease Control Priorities study and the GBD project both used a 3% discount rate, and the US Panel on Cost-Effectiveness in Health and Medicine recently recommended that economic analyses of health also use a 3% real discount rate to adjust both costs and health outcomes [42].

The YLL is estimated as:

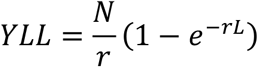

where, N= number of deaths

L= Life expectancy at age of death

r= discount rate (we have also used 3% discount rate)

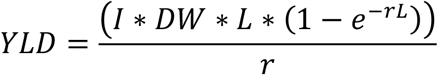

where, I= number of incidence/prevalence cases. For acute diseases, incidence is considered same as prevalence

DW= disability weight (a weight factor that reflects the severity of the disease on a scale from 0 (perfect health) to 1 (dead)

L= duration of disability

r= discount rate

As COVID-19 is a novel disease, its disability weight is not available. Since COVID-19 is a severe infectious disease having acute period, we have used the disability weight of 0.133 for Infectious disease (acute episode, severe category) as proxy for COVID-19 [43]. The duration of disability of 60 days is used because the patients of COVID-19 have been hospitalized for on average 30 days and after discharge and quarantined for 14-28 days approximately.

## Results

Table 1 presents the key indicators of COVID-19 attributable mortality in four countries under study. With additional 93,858 deaths in USA, 32,169 deaths in Italy, 8213 deaths in Germany and 3925 deaths in Sweden in a span of about three months, the share of COVID-19 deaths amounts to 3.2% of total deaths in USA, 4.8% in Italy, 0.9% in Germany and 4.1% in Sweden. The COVID-19 attributable deaths can be considered as additional deaths avoidable without this infection. The case fatality ratio was very high in Italy (14.2) followed by Sweden (11.9) and USA (5.6). The pandemic has infected at least 0.5% of the population in USA, 0.4% in Italy, 0.3% in Sweden and 0.2% in Germany. The COVID-19 attributable deaths has already lower life expectancy by 0.4 years for USA and Sweden, 0.5 years in Italy and 0.1 years in Germany. At 10% share, the reduction in life expectancy would be 1.2 years for USA, 1 years for both Italy and Germany and 0.9 years for Sweden (Appendix 1).

**Table 1:** **Summary indicators of population and COVID-19 attributable mortality indicators in USA, Italy, Germany and Sweden, 2020**

| Country | USA | Italy | Germany | Sweden |
| --- | --- | --- | --- | --- |
| Total Population in million (2020) | 331.0 | 60.5 | 83.8 | 10.1 |
| Estimated number of annual deaths without COVID-19 (2020) | 2,830,600 | 633,800 | 923,800 | 91,200 |
| Number of deaths with COVID-19 (As of 20th May) | 93,858 | 32,169 | 8,213 | 3,925 |
| Total Deaths including deaths due to COVID-19 | 2,924,458 | 665,969 | 932,013 | 95,125 |
| COVID-19 death as a share of total deaths as of 20th May 2020 | 3.21 | 4.83 | 0.88 | 4.13 |
| Case-fatality ratio | 5.95 | 14.19 | 4.61 | 11.96 |
| Total number of COVID-19 infection as of 20th May | 1,576,950 | 226,699 | 178,170 | 32,809 |
| Infection rate | 0.48 | 0.37 | 0.21 | 0.32 |
| Estimated Life expectancy without COVID-19 (years) | 79.5 | 83.6 | 81.5 | 82.7 |
| Reduction in life expectancy without actual number COVID-19 deaths accounting to 3.2% (in Years) | 0.4 | 0.5 | 0.1 | 0.4 |
| Reduction in life expectancy without actual number COVID-19 deaths accounting to 6% (in Years) | 0.7 | 0.6 | 0.6 | 0.5 |
| Reduction in life expectancy without actual number COVID-19 deaths accounting to 10% (in Years) | 1.2 | 1.0 | 1.0 | 0.9 |

Appendix 2(a) presents life table probability of death with and without COVID-19 in Italy, Germany and Sweden and Appendix 2(b) presents the same for USA (due to dissimilarity in age group). The probability of death without COVID-19 was lowest in Sweden followed by Italy and Germany. The COVID-19 attributable deaths have disproportionately increased the probability of death in 70+ age group in all these four countries. The pattern is also similar for USA.

Estimates from life table with and without COVID-19 for these four countries exhibit the changing age-specific survival patterns (Table 2, Appendix 3). The life expectancy for 2020 was 79.5 years in USA, 83.6 years in Italy, 81.5 years in Germany and 82.7 years in Sweden. The additional deaths due to COVID-19 results in a rise in CDR from 10.5 to 11 in Italy and this would rise to 11.6 with the COVID-19 death share rising to10%. In case of USA, it has also increased from 8.6 to 9.0 and the pattern is similar in Germany and Sweden as well.

**Table 2:** **Life expectancy under alternative scenarios of COVID-19 attributable mortality in United States of America (USA), 2020**

| Age Group | LE without COVID19 | LE with actual number of COVID-19 deaths (3.2%) | LE with COVID-19 accounting 6% of total deaths | LE with COVID-19 accounting 10% of total deaths |
| --- | --- | --- | --- | --- |
| 0-1 | 79.5 | 79.1 | 78.7 | 78.3 |
| 1-4 | 78.8 | 78.4 | 78.1 | 77.6 |
| 5-14 | 74.9 | 74.5 | 74.2 | 73.7 |
| 15-24 | 65.0 | 64.6 | 64.3 | 63.8 |
| 25-34 | 55.5 | 55.1 | 54.8 | 54.3 |
| 35-44 | 46.2 | 45.8 | 45.4 | 45.0 |
| 45-54 | 37.0 | 36.6 | 36.2 | 35.8 |
| 55-64 | 28.3 | 28.0 | 27.6 | 27.2 |
| 65-74 | 20.5 | 20.1 | 19.9 | 19.5 |
| 75-84 | 13.3 | 13.0 | 12.7 | 12.4 |
| 85+ | 7.6 | 7.4 | 7.2 | 6.9 |

Note: LE- Life expectancy

Age specific assessment of Years of potential life lost (YPLL) under varying scenario of COVID-19 death share is presented in Table 3 and Appendix 4. While YPLL without COVID-19 was 55.2 million in USA, 8.9 million in Italy, 14.4 million in Germany and 1.3 million in Sweden, COVID-19 has added 1.5 million, 0.5 million, 0.1 million and 0.04 millions of YPLL in USA, Italy, Germany, and Sweden, respectively. Rate of YPLL (per 1000n population) is highest in Italy (7.4) followed by USA (4.7) and Sweden (4.6). With rising share of COVID-19 deaths to the tune of 6% and 10%, The share of YPLL on this count will rise from 4.7 to 8.6 and 14.1 per 1000 population, respectively in USA. Similar pattern has been observed for Italy, Germany and Sweden. Higher age-groups (45 years and above) are contributing more than 70% of YPLL in all the countries.

**Table 3:** **Years of Potential Life Lost (YPLL) under varying scenario of COVID-19 attributable deaths in USA, 2020**

| Years of Potential Life Lost (YPLL) |  |  |  |  |  |  |
| --- | --- | --- | --- | --- | --- | --- |
| Age Group | Deaths from all cause Without COVID-19 | COVID-19 deaths as of 20<sup>th</sup> May,2020 | COVID-19 deaths accounting 6% death share | COVID-19 deaths accounting 10% death share | Share of YPLL without COVID-19 deaths | Share of YPLL with COVID-19 deaths as of 20<sup>th</sup> May, 2020 |
| 0-1 | 1597660 | 323 | 601 | 995 | 2.89 | 0.02 |
| 1-4 | 528026 | 213 | 397 | 658 | 0.96 | 0.01 |
| 5-14 | 404728 | 710 | 1321 | 2188 | 0.73 | 0.05 |
| 15-24 | 2198402 | 6682 | 12426 | 20560 | 3.98 | 0.43 |
| 25-34 | 3408302 | 34703 | 64475 | 106542 | 6.17 | 2.24 |
| 35-44 | 3730989 | 73836 | 137011 | 226015 | 6.76 | 4.76 |
| 45-54 | 6286116 | 166082 | 307663 | 506338 | 11.38 | 10.72 |
| 55-64 | 10504149 | 316168 | 584397 | 958838 | 19.02 | 20.40 |
| 65-74 | 10931986 | 395769 | 729310 | 1191610 | 19.80 | 25.54 |
| 75-84 | 8940175 | 328953 | 603261 | 979131 | 16.19 | 21.23 |
| 85+ | 6686000 | 226234 | 410856 | 657812 | 12.11 | 14.60 |
| **Total** | **55216531** | **1549673** | **2851718** | **4650687** | **100.00** | **100.00** |
| **Rate of YPLL** | **166.8** | **4.7** | **8.6** | **14.1** | **NA** | **NA** |

The estimated DALY at current share of attributable COVID-19 deaths is 1.17 million in USA, 0.35 million in Italy, 0.08 million in Germany and 0.03 million in Sweden. At 6% share of attributable COVID-19 deaths, DALY for all ages would be 2.19 million in USA, 0.44 million in Italy, 0.59 million in Germany and 0.05 million in Sweden. The COVID-19 attributed loss has increased the DALYs by 5.86 per 1000 population in Italy, 3.5 in USA, 1.04 in Germany and 0.45 in Sweden (Table 4 & Appendix 5). If COVID-19 attributable death accounts 6% death share, the DALYs would be 7.28 per 1000 population in Italy, 6.6 in USA 7.10 in Germany and 0.66 in Sweden. Similarly, when COVID-19 accounts 10% death share, DALYs is 12.13 per 1000 population in Italy, 11.0 in USA, 11.83 in Germany and 1.09 in Sweden. Among all the four countries, the population 70 years and above account more than three-fourth contribution in DALY while younger ages have relatively low contribution in all the scenarios.

**Table 4:** **Estimate of DALY with varying scenario of COVID-19 in USA, 2020**

| Age Group | DALY |  |  | DALY per 1000 population |  |  |
| --- | --- | --- | --- | --- | --- | --- |
|  | COVID-19 deaths as of 20<sup>th</sup> May,2020 | COVID-19 deaths accounting 6% death share | COVID-19 deaths accounting 10% death share | COVID-19 deaths as of 20<sup>th</sup> May,2020 | COVID-19 deaths accounting 6% death share | COVID-19 deaths accounting 10% death share |
| 0-1 | 131 | 245 | 408 | 0.0 | 0.0 | 0.1 |
| 1-4 | 87 | 163 | 272 | 0.0 | 0.0 | 0.0 |
| 5-14 | 301 | 564 | 939 | 0.0 | 0.0 | 0.0 |
| 15-24 | 3147 | 5883 | 9805 | 0.1 | 0.1 | 0.2 |
| 25-34 | 18184 | 33995 | 56658 | 0.4 | 0.7 | 1.2 |
| 35-44 | 41805 | 78155 | 130259 | 1.0 | 1.9 | 3.1 |
| 45-54 | 102728 | 192051 | 320084 | 2.5 | 4.7 | 7.9 |
| 55-64 | 217011 | 405702 | 676170 | 5.1 | 9.6 | 16.0 |
| 65-74 | 301684 | 563999 | 939998 | 9.4 | 17.5 | 29.2 |
| 75-84 | 278222 | 520136 | 866894 | 17.2 | 32.1 | 53.5 |
| 85+ | 210036 | 392663 | 654438 | 31.4 | 58.7 | 97.9 |
| **Total** | **1173338** | **2193555** | **3655925** | **3.5** | **6.6** | **11.0** |

## Discussion and Conclusion

The COVID-19 pandemic is one of the worst ever misery posed to mankind. While epidemics in the past have gripped limited geographical boundaries, the COVID-19 has engulfed the entire world within a brief period of four months with a reasonable degree of spread potential. Apart from threat to human life, its containment measures have led to economic loss and generated psychological scare among individuals, households, community and the nation at large. The COVID-19 pandemic has paralysed the economic activities, deepened the global recession and has assumed a crisis proportion worldwide. Given the scale and intensity of this pandemic, this is first attempt in our knowledge to assess the mortality attributed to COVID-19 in four worst affected countries. Such an assessment involves the extent of reduction in life expectancy, person year life lost and DALY that are yet to be made available so far. Selection of countries are primarily based on the extent of severity of the pandemic and availability of data but the exercise can very well be replicated elsewhere. The following are the salient findings.

First and foremost, COVID19 induced fatalities have undoubtedly contributed towards rise in the overall mortality rate in all four countries. The death rate has increased from 9.0 without COVID-19 to 9.4 with COVID-19 in USA, from 11.0 to 11.6 in Italy, 11.0 to 11.1 in Germany and 9.1 to 10.0 in Sweden. Second, the life expectancy has compressed by 0.4 years each in USA and Sweden, 0.5 years in Italy and 0.1 years in Germany. Within a few months, the COVID-19 attributable death share amounts to about 3% in USA, 4% in Sweden and 5% in Italy. If this trend of mortality continues till end of the year, reduction in life expectancy would be substantial in these countries. Third, most of the COVID-19 deaths are unwarranted, untimely and premature. COVID-19 attributable deaths have already added 1.5 million, 0.5 million, 0.1 million and 0.04 millions of YPLL in USA, Italy, Germany, and Sweden, respectively. Fourth, with less than 1% infection, the DALY a from COVID-19 was 3.5 per thousand in USA, 5.86 in Italy, 1.04 in Germany and 0.45 in Sweden. If the spread of COVID-19 goes unabated, the loss of DALY would be similar to high fatality disease.

These findings are markers of tragedy experienced in countries ranked high in the level of human development, higher income level and are said to be having a better health care system. Hence the failure of preparedness to confront this pandemic by the developed world exposes our vulnerability to emerging infection of similar kind in future. In the absence of a vaccine as well as no systematic medical intervention, the only way out is the containment of its spread or developing a herd immunity in due course. At present great efforts are made by national and local government for management and control of pandemic by diverting the resources (financial and physical) for health care and lock down measures.

We acknowledge that the COVID-19 attributable deaths are to some extent underestimated due to lack of comprehensive testing, under-reporting and misclassification of COVID-19 deaths in these countries. Despite these limitations, these estimates of mortality pattern do signals about its long-term implications towards structural and compositional balance of population across world regions. Though it is very early to gauge its final impact on population structure and composition, its persistence with its virulence unless curbed by introduction of an effective vaccine and means of cure may well change the world order to a significant extent.

## Data Availability

The data from United Nation Population Projection, worldmeter.info, Statista and Centre for disease control and Prevention has been collected for analyses.

https://population.un.org/wpp/DataQuery/

https://data.cdc.gov/NCHS/Provisional-COVID-19-Death-Counts-by-Sex-Age-and-S/9bhg-hcku/data

https://www.statista.com/statistics/1105061/coronavirus-deaths-by-region-in-italy/

https://www.statista.com/statistics/1105512/coronavirus-covid-19-deaths-by-gender-germany).

https://www.statista.com/statistics/1107913/number-of-coronavirus-deaths-in-sweden-by-age-groups;

https://www.worldometers.info/coronavirus/

## Appendix

**Appendix 1:**
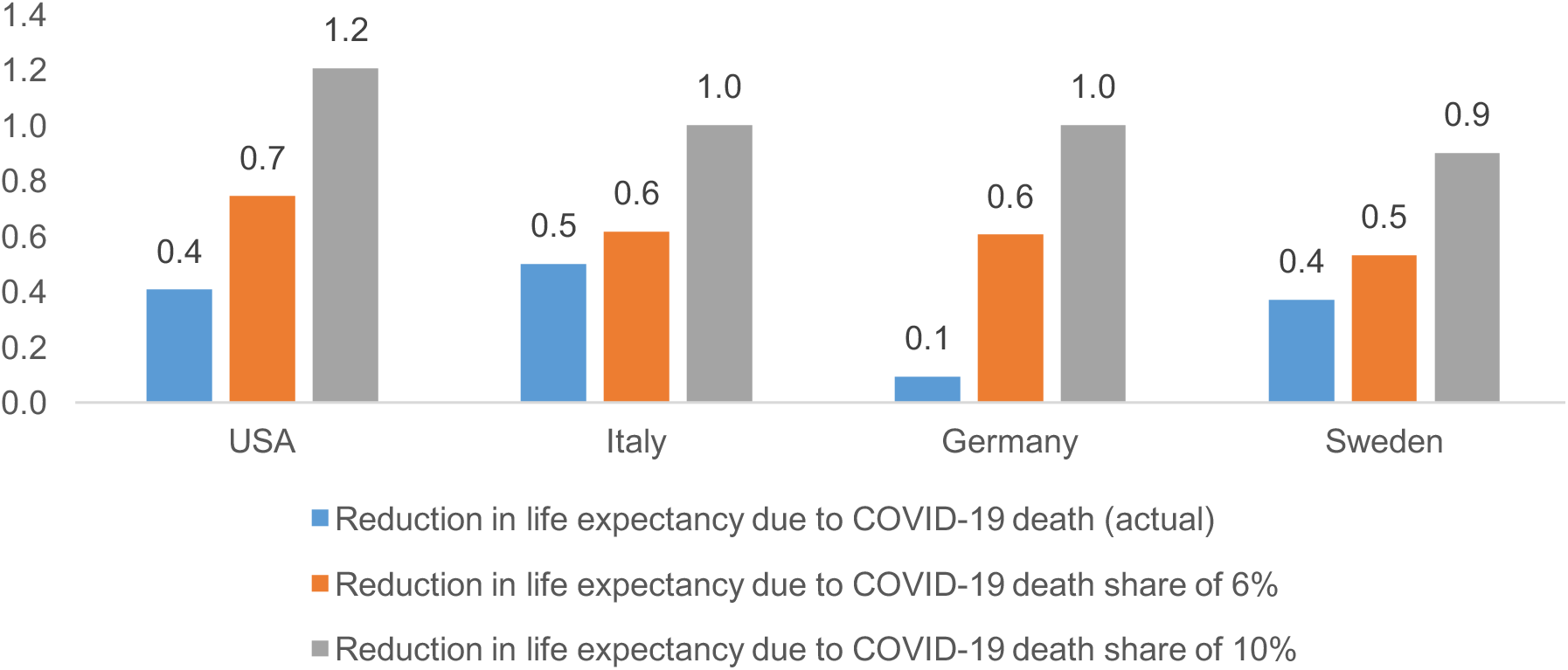
Reduction in Life Expectancy at Birth (years) due to COVID-19 attributable deaths in USA, Italy, Germany and Sweden, 2020.

**Appendix 2(a):** **Life table Probability of death without and with COVID-19 attributable deaths in Italy, Germany and Sweden, 2020**

| Age group | Italy |  | Germany |  | Sweden |  |
| --- | --- | --- | --- | --- | --- | --- |
|  | Probability at death without COVID-19 | Probability of death with COVID-19 death share of 4.8 % (actual) | Probability at death without COVID-19 | Probability of death with COVID-19 death share of 0.9% (actual) | Probability at death without COVID-19 | Probability of death with COVID-19 death share of 4.1 % (actual) |
| 0-9 | 0.0032 | 0.0032 | 0.0041 | 0.0041 | 0.0017 | 0.0017 |
| 10-19 | 0.0014 | 0.0014 | 0.0018 | 0.0018 | 0.0018 | 0.0018 |
| 20-29 | 0.0029 | 0.0030 | 0.0036 | 0.0036 | 0.0047 | 0.0047 |
| 30-39 | 0.0046 | 0.0047 | 0.0059 | 0.0059 | 0.0060 | 0.0061 |
| 40-49 | 0.0121 | 0.0124 | 0.0190 | 0.0190 | 0.0126 | 0.0129 |
| 50-59 | 0.0286 | 0.0298 | 0.0441 | 0.0443 | 0.0304 | 0.0313 |
| 60-69 | 0.0729 | 0.0770 | 0.0930 | 0.0936 | 0.0824 | 0.0848 |
| 70-79 | 0.1829 | 0.1950 | 0.2550 | 0.2569 | 0.1845 | 0.1917 |
| 80-89 | 0.5118 | 0.5312 | 0.5248 | 0.5289 | 0.5381 | 0.5577 |

**Appendix 2(b):**
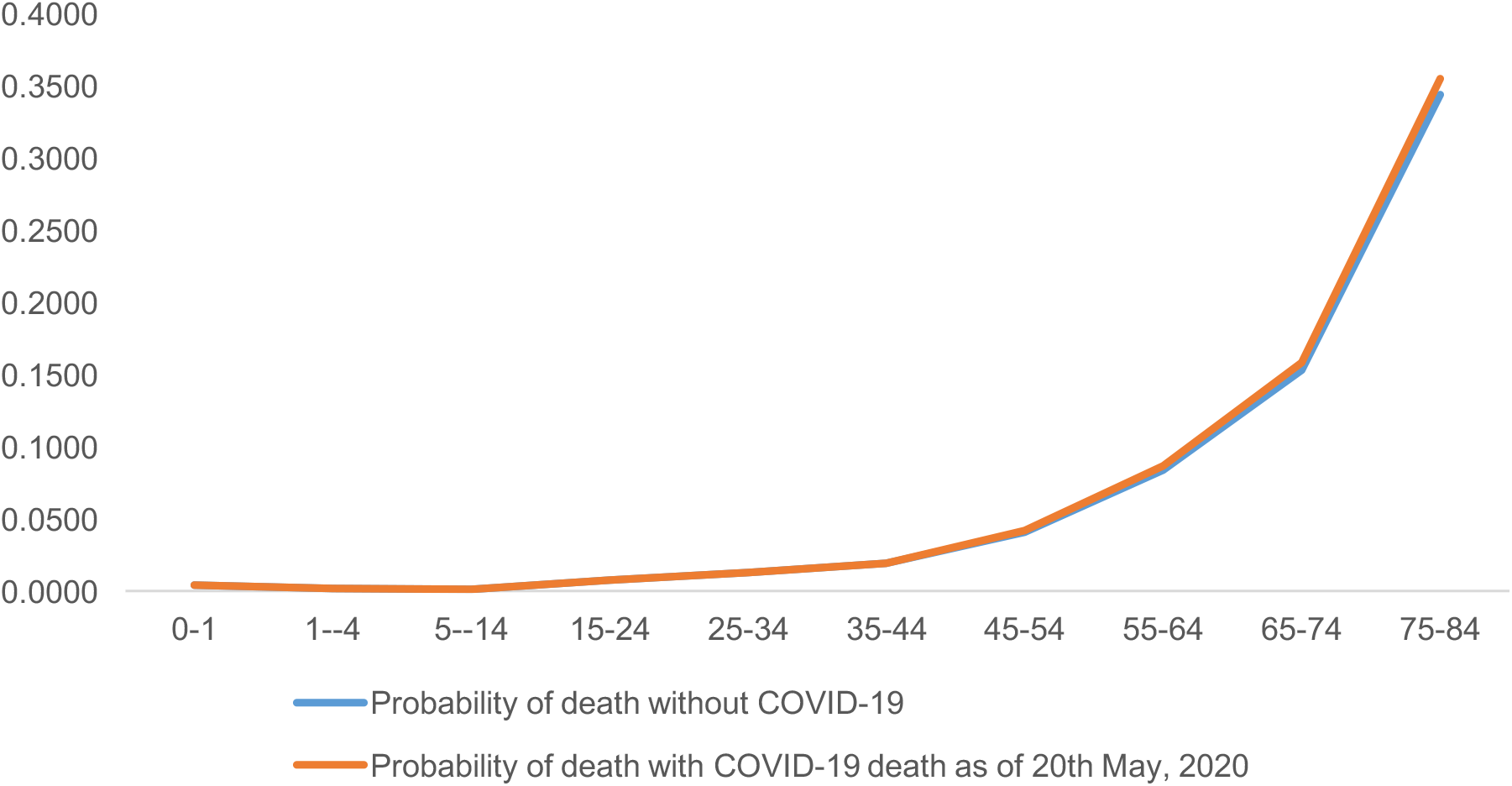
Probability of death with and without COVID-19 in USA, 2020.

**Appendix 3:**
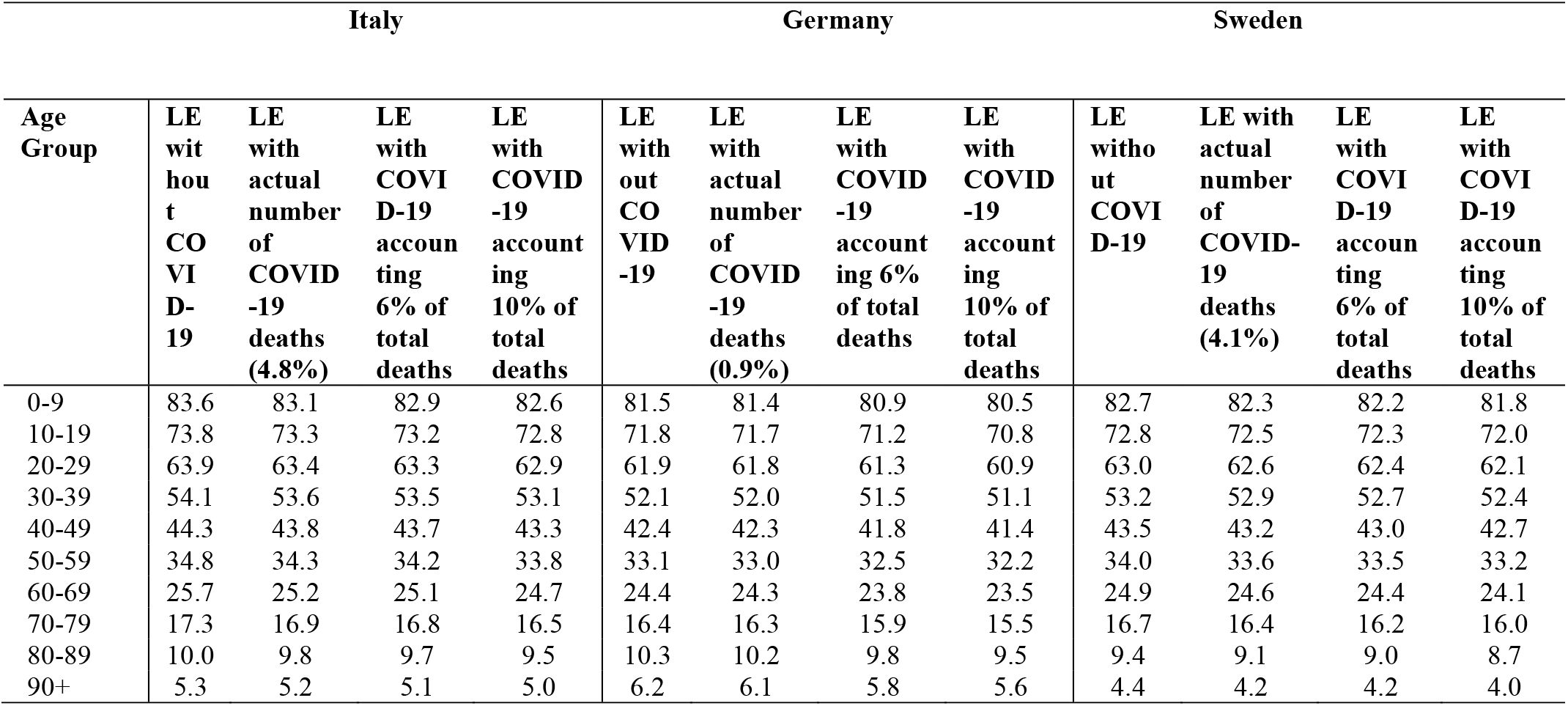
**Life expectancy under alternative scenarios of COVID-19 attributable mortality in Italy, Germany and Sweden, 2020**

**Appendix 4:**
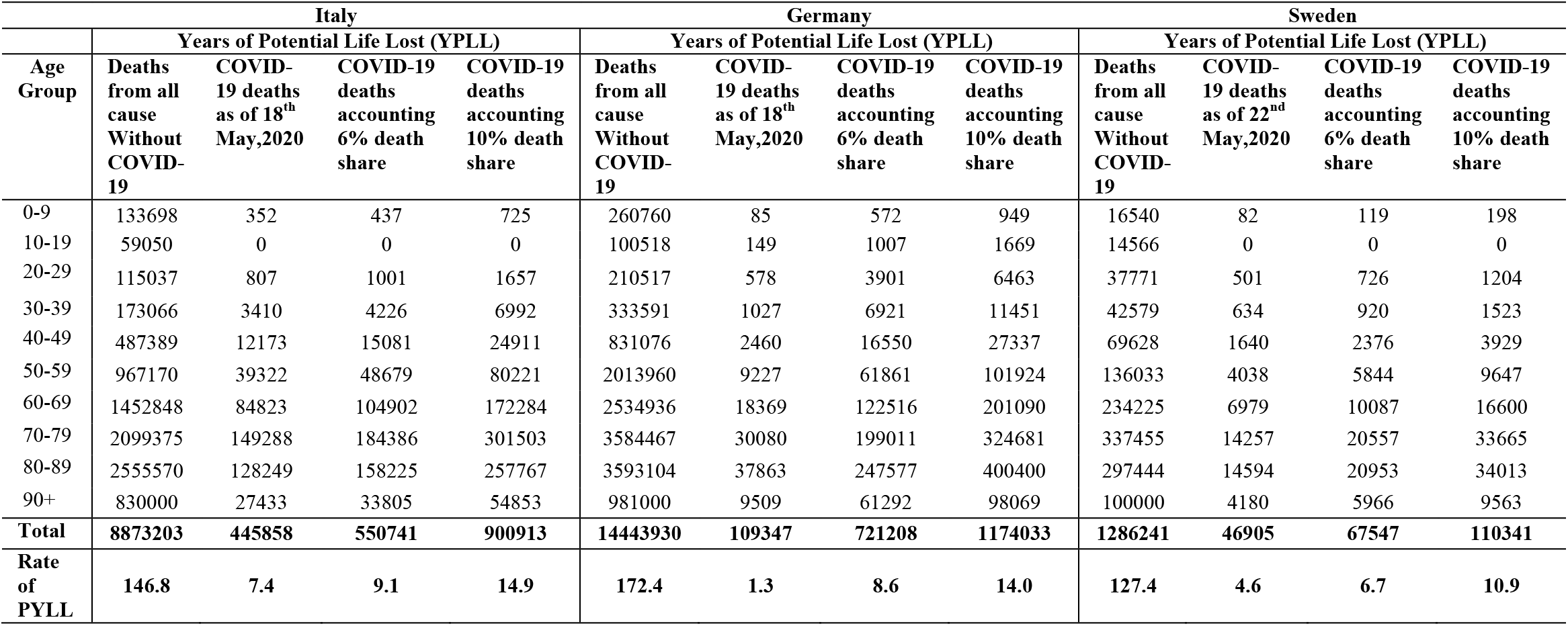
**Years of Potential Life Lost (YPLL) under varying scenario of COVID-19 attributable deaths in Italy, Germany and Sweden, 2020**

**Appendix 5:**
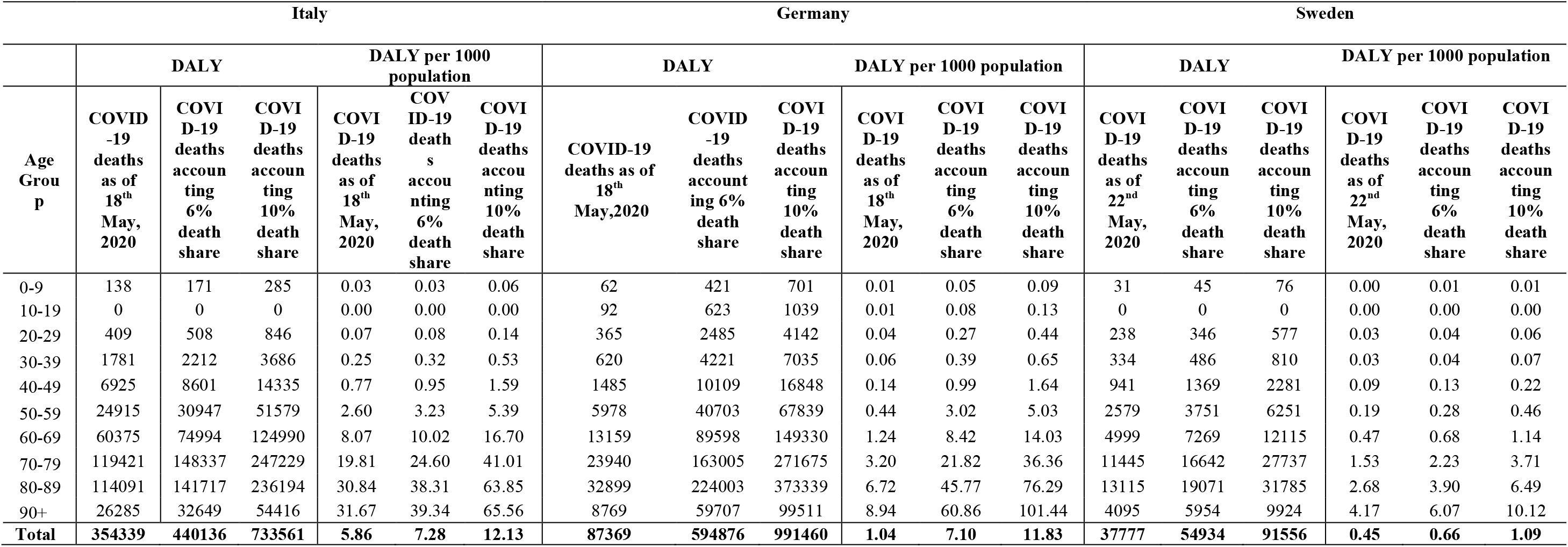
**Estimate of DALY with varying scenario of COVID-19 in Italy, Germany and Sweden, 2020**

## References

[1] Baud, D., Qi, X., Nielsen-Saines, K., Musso, D., Pomar, L., & Favre, G. (2020). Real estimates of mortality following COVID-19 infection. The Lancet infectious diseases.

[2] Leung, C. (2020). Risk factors for predicting mortality in elderly patients with COVID-19: a review of clinical data in China. Mechanisms of Ageing and Development, 111255.

[3] Steyn, N., Binny, R. N., Hannah, K., Hendy, S., James, A., Kukutai, T., Lustig, A., McLeod, M., Plank, M. J., Ridingd, K., & Sporle, A. (2020). Estimated inequities in COVID-19 infection fatality rates by ethnicity for Aotearoa New Zealand. medRxiv.

[4] Onder, G., Rezza, G., & Brusaferro, S. (2020). Case-fatality rate and characteristics of patients dying in relation to COVID-19 in Italy. Jama, 323(18), 1775–1776.

[5] Verity, R., Okell, L. C., Dorigatti, I., Winskill, P., Whittaker, C., Imai, N., Dannenburg, G. C., Thompson, H., Walker, P. G., Fu, H., & Dighe, A. (2020). Estimates of the severity of coronavirus disease 2019: a model-based analysis. The Lancet Infectious Diseases.

[6] Lai, C. C., Wang, C. Y., Wang, Y. H., Hsueh, S. C., Ko, W. C., & Hsueh, P. R. (2020). Global epidemiology of coronavirus disease 2019: disease incidence, daily cumulative index, mortality, and their association with country healthcare resources and economic status. International journal of antimicrobial agents, 105946.

[7] Mahase, E. (2020). Covid-19: death rate is 0.66% and increases with age, study estimates. BMJ (Clinical research ed.), 369, m1327.

[8] Dowd, J. B., Andriano, L., Brazel, D. M., Rotondi, V., Block, P., Ding, X., Liu, Y & Mills, M. C. (2020). Demographic science aids in understanding the spread and fatality rates of COVID-19. Proceedings of the National Academy of Sciences.

[9] Chang, D., Lin, M., Wei, L., Xie, L., Zhu, G., Cruz, C. S. D., & Sharma, L. (2020). Epidemiologic and clinical characteristics of novel coronavirus infections involving 13 patients outside Wuhan, China. Jama, 323(11), 1092–1093.

[10] Shen, K. L., & Yang, Y. H. (2020). Diagnosis and treatment of 2019 novel coronavirus infection in children: a pressing issue.

[11] Wang, D., Hu, B., Hu, C., Zhu, F., Liu, X., Zhang, J., Wang, B., Xiang, H., Cheng, Z., Xiong, Y., & Zhao, Y. (2020). Clinical characteristics of 138 hospitalized patients with 2019 novel coronavirus–infected pneumonia in Wuhan, China. Jama, 323(11), 1061–1069.

[12] Chen, N., Zhou, M., Dong, X., Qu, J., Gong, F., Han, Y., Qiu, Y., Wang, J., Liu, Y., Wei, Y., Xia, J., & Yu, T. (2020). Epidemiological and clinical characteristics of 99 cases of 2019 novel coronavirus pneumonia in Wuhan, China: a descriptive study. The Lancet, 395(10223), 507–513.

[13] Ji, Y., Ma, Z., Peppelenbosch, M. P., & Pan, Q. (2020). Potential association between COVID-19 mortality and health-care resource availability. The Lancet Global Health, 8(4), e480.

[14] Dilcher, M., Werno, A., & Jennings, L. C. (2020). SARS-CoV-2: a novel deadly virus in a globalised world. NZ Med J, 133, 6–11.

[15] Zhou, F., Yu, T., Du, R., Fan, G., Liu, Y., Liu, Z., Xiang, J., Wang, Y., Song, B., Gu, X., & Guan, L. (2020). Clinical course and risk factors for mortality of adult inpatients with COVID-19 in Wuhan, China: a retrospective cohort study. The lancet.

[16] Chow, N., Fleming-Dutra, K., Gierke, R., Hall, A., Hughes, M., Pilishvili, T., Ritchey, M., Roguski, K., Skoff, T. and Ussery, E., 2020. Preliminary estimates of the prevalence of selected underlying health conditions among patients with coronavirus disease 2019—United States, February 12–March 28, 2020.

[17] Bonow, R. O., Fonarow, G. C., O’Gara, P. T., & Yancy, C. W. (2020). Association of coronavirus disease 2019 (COVID-19) with myocardial injury and mortality. JAMA cardiology.

[18] Lippi, G., Wong, J., & Henry, B. M. (2020). Hypertension and its severity or mortality in Coronavirus Disease 2019 (COVID-19): a pooled analysis. Polish archives of internal medicine.

[19] Pal, R., & Bhadada, S. K. (2020). COVID-19 and non-communicable diseases. Postgraduate Medical Journal.

[20] Madjid, M., Safavi-Naeini, P., Solomon, S. D., & Vardeny, O. (2020). Potential effects of coronaviruses on the cardiovascular system: a review. JAMA cardiology.

[21] Lei, S., Jiang, F., Su, W., Chen, C., Chen, J., Mei, W., Zhan, L. Y., Jia, Y., Zhang, L., Liu, D., & Xia, Z. Y. (2020). Clinical characteristics and outcomes of patients undergoing surgeries during the incubation period of COVID-19 infection. EClinicalMedicine, 100331.

[22] Guan, W. J., Ni, Z. Y., Hu, Y., Liang, W. H., Ou, C. Q., He, J. X., … & Du, B. (2020). Clinical characteristics of coronavirus disease 2019 in China. New England journal of medicine, 382(18), 1708–1720.

[23] Russell, T. W., Hellewell, J., Jarvis, C. I., Van Zandvoort, K., Abbott, S., Ratnayake, R., Flasche, S., Eggo, R. M., Edmunds, W. J., Kucharski, A. J., & CMMID COVID-19 working group. (2020). Estimating the infection and case fatality ratio for coronavirus disease (COVID-19) using age-adjusted data from the outbreak on the Diamond Princess cruise ship, February 2020. Eurosurveillance, 25(12), 2000256.

[24] Weinberger, D., Cohen, T., Crawford, F., Mostashari, F., Olson, D., Pitzer, V. E., Reich, N. G., BS, M. R., Simonsen, L., Watkins, A., & Viboud, C. (2020). Estimating the early death toll of COVID-19 in the United States. Medrxiv.

[25] Mohanty and Sahoo (2020). COVID-19 and Mortality: India’s Perspective, Working paper No 11, IIPS Analytical Series on Covid 19, IIPS, Mumbai.

[26] Ghislandi, S., Muttarak, R., Sauerberg, M., & Scotti, B. (2020). News from the front: Estimation of excess mortality and life expectancy in the major epicenters of the COVID-19 pandemic in Italy. medRxiv.

[27] Goldstein, J. R., & Lee, R. D. (2020). Demographic Perspectives on Mortality of Covid-19 and Other Epidemics (No. w27043). National Bureau of Economic Research.

[28] Fielding-Miller, R. K., Sundaram, M. E., & Brouwer, K. (2020). Social determinants of COVID-19 mortality at the county level. medRxiv.

[29] Kulu, H., & Dorey, P. (2020). The contribution of age structure to the number of deaths from Covid-19 in the UK by geographical units. medRxiv.

[30] Guilmoto, C. Z. (2020). COVID-19 death rates by age and sex and the resulting mortality vulnerability of countries and regions in the world. medRxiv.

[31] Banerjee, A., Pasea, L., Harris, S., Gonzalez-Izquierdo, A., Torralbo, A., Shallcross, L., Noursadeghi, M., Pillay, D., Sebire, N., Holmes, C., & Pagel, C. (2020). Estimating excess 1-year mortality associated with the COVID-19 pandemic according to underlying conditions and age: a population-based cohort study. The Lancet.

[32] Dudel, C., Riffe, T., Acosta, E., van Raalte, A. A., & Myrskyla, M. (2020). Monitoring trends and differences in COVID-19 case fatality rates using decomposition methods: Contributions of age structure and age-specific fatality. medRxiv.

[33] Bohk-Ewald, C., Dudel, C., & Myrskylä, M. (2020). A demographic scaling model for estimating the total number of COVID-19 infections. arXiv preprint arXiv:2004.12836.

[34] https://population.un.org/wpp/DataQuery/Accessed on 20thMay 2020

[35] https://data.cdc.gov/NCHS/Provisional-COVID-19-Death-Counts-by-Sex-Age-and-S/9bhg-hcku/data

[36] https://www.statista.com/statistics/1105061/coronavirus-deaths-by-region-in-italy/statista.com/

[37] statistics/1105512/coronavirus-covid-19-deaths-by-gender-germany).

[38] statista.com/statistics/1107913/number-of-coronavirus-deaths-in-sweden-by-age-groups;

[39] https://www.worldometers.info/coronavirus/

[40] Gardner, J. W., & Sanborn, J. S. (1990). Years of potential life lost (YPLL)—what does it measure? Epidemiology, 322–329.

[41] Werber, D., Hille, K., Frank, C., Dehnert, M., Altmann, D., Müller-Nordhorn, J., Koch, J., & Stark, K. (2013). Years of potential life lost for six major enteric pathogens, Germany, 2004–2008. Epidemiology & Infection, 141(5), 961–968.

[42] Gold, M. R., Stevenson, D., & Fryback, D. G. (2002). HALYS and QALYS and DALYS, Oh My: similarities and differences in summary measures of population Health. Annual review of public health, 23(1), 115–134.

[43] Salomon JA, Haagsma JA, Davis A, et al. Disability weights for the Global Burden of Disease 2013 study. Lancet Glob Health 2015;3:e712–723. doi:10.1016/S2214-109X(15)00069-8

